# Impact of Meteorological factors and population size on the transmission of Micro-size respiratory droplets based Coronavirus: A brief study of highly infected cities in Pakistan

**DOI:** 10.1101/2020.07.14.20153544

**Authors:** Iram Shahzadi, Anum Shahzadi, Junaid Haider, Sadia Naz, Rai. M. Aamir, Ali Haider, Hafiz Rizwan Sharif, Imran Mahmood Khan, Muhammad Ikram

**Author notes:** **Corresponding author:** Sadia Naz, Tianjin Institute of Industrial Biotechnology, Chinese Academy of Sciences, Tianjin 300308, China., E-mail address, Muhammad Ikram, Solar Cell Applications Research Lab, Department of Physics, Government College University Lahore, 54000, Punjab, Paksitan.

## Abstract

Ongoing Coronavirus epidemic (COVID-19) identified first in Wuhan, China posed huge impact on public health and economy around the globe. Both cough and sneeze based droplets or aerosols encapsulated COVID-19 particles are responsible for air borne transmission of this virus and caused unexpected escalation and high mortality worldwide. Current study intends to investigate correlation of COVID-19 epidemic with meteorological parameters particularly, temperature, rainfall, humidity, and wind speed along with population size. Data set of COVID-19 for highly infected cities of Pakistan was collected from the official website of National Institute of health (NIH). Spearman’s rank (r*s*) correlation coefficient test employed for data analysis revealed significant correlation between temperature minimum (TM), temperature average (TA), wind speed (WS) and population size (PS) with COVID-19 pandemic. Furthermore, receiver operating characteristics (ROC) curve was used to analyze the sensitivity of TA, WS, and PS on transmission rate of COVID-19 in selected cities of Pakistan. The results obtained for sensitivity and specificity analysis for all selected parameters signifies sensitivity and direct correlation of COVID-19 transmission with temperature variation, WS and PS. Positive correlation and strong association of PS parameter with COVID-19 pandemic suggested need of more strict actions and control measures for highly populated cities. These findings will be helpful for health regulatory authorities and policymakers to take specific measures to combat COVID-19 epidemic in Pakistan.

## 1. Introduction

Recent outbreak of coronavirus disease characterized by pneumonia of unexplained etiology was first reported from Wuhan, China in December, 2019 that later spread globally (Li et al., 2020; Zhu et al., 2020). Coronavirus (COVID-19, or 2019-nCoV) epidemic posed huge impact on public health and economy all over the world and has been declared as public health emergency by World Health Organization (WHO) on January 30, 2020 (Rothan and Byrareddy, 2020). Later, International Committee on Taxonomy of Viruses named this novel coronavirus as severe acute respiratory syndrome coronavirus 2 (SARS-CoV-2) on 11^th^ February 2020 (Yang and Wang, 2020). The COVID-19 has more severe effects than Severe Acute Respiratory Syndrome (SARS) and Middle East Respiratory Syndrome (MERS), although phylogenetically similar (Van Doremalen et al., 2013). Common symptoms of this pandemic include fever, cough, troubled breathing, fatigue, body pain and lesions on patient’s lungs (Guan et al., 2020; Song et al., 2020). In severe conditions, patients suffer from viral pneumonia, diarrhea, acute cardiac injury and RNAaemia (Huang et al., 2020). Nevertheless, recently few cases of corona virus have been claimed by Chinese government with no observed clinical symptoms (asymptomatic) of COVID-19 on May 18, 2020.

This pandemic has wrapped whole globe and effected 216 countries including Pakistan. In Pakistan, first 2 cases of Corona virus were confirmed by Federal Health Minister of Pakistan on Feb 26, 2020 in Islamabad and Karachi that arrived to twenty (14 in Sindh, 5 in Gilgit Baltistan, 1 in Baluchistan in 12 yr. old boy) till March 12, 2020. History of these patients depicts their recent visit to Iran, London or Syria (Ali, 2020; Saqlain et al., 2020). Pakistan share borders with countries infected with COVID-19 namely, China, and Iran (Remuzzi and Remuzzi, 2020; Zhu et al., 2020). First coronavirus death from Pakistan was reported in Peshawar on March 18, 2020. Up to now, a total 1,372,825 tests, 225,282 confirmed cases and 4,619 deaths have been reported by NIH, and more than 54% of total confirmed cases of Pakistan are reported from three highly infected cities such as Karachi (70,143 cases), Lahore (41,416 cases) and Peshawar (11,134 cases) till July 4, 2020. To control this pandemic, the government of Pakistan has taken strict actions like screening of passengers traveling from other countries, restrict inter-city transportation, early detection of new cases, quarantine and restrict people mobility (Noreen et al.). In addition, people are advised to use facemasks, hand sanitizers and maintain a distance of one meter to avoid transmission of virus by direct contact (Organization, 2020; Saqlain et al., 2020).

In fact, spread of SARS-CoV-2 is attributed to three different routes of its transmission i.e. (a) Aerosol transmission in confined areas, (b) direct inhalation of large respiratory droplets with diameter ranged 5-10 μm produced by cough or sneeze, (c) Direct contact with surfaces contaminated with virus (Lipsitch et al., 2020). Both, droplet and aerosol are two major routes for transmission of COVID-19 where viral particles spread through breath, sneeze and cough of an infected person. Droplets are large sized mucus or saliva globs (size > 5μm) with virus encapsulated inside that fall in close proximity of their origin while aerosols are comparatively small sized particles (size < 5μm) that transmit to larger distance and may lead to higher rate of COVID-19 spread (Grayson et al., 2017; Liu et al., 2017), as depicted in Figure 1. Both cough and sneeze emit viral particles in the form of aerosol droplets where cough based particles having diameter < 20 μm quickly loss water and achieve diameter almost less than half of initial size (Nicas et al., 2005). Aerosol can survive in air for long time and can penetrate into alveolar region of lungs causing viral infection deep in alveolar tissues (Tellier, 2009). The distance travel by sneeze and cough based viral particles depends on the speed of droplet clouds, air flow, temperature and humidity of air i.e. larger droplets travel up to 2m (speed 10m/s) from initial point and up to 1m if emitted at speed of 1m/s while sneeze based droplets travel up to 7-8m (Xie et al., 2007). In addition, Bioaerosol also known as droplet nuclei have COVID-19 encapsulated inside particles released from expiratory activities of infected person with size in range of 4 to 8 μm (i.e. 95% < 100 μm) and their viability lasts for 3 h (Jayaweera et al., 2020). Similarly, droplets of COVID-19 showed trend similar to other SARS viruses in terms of stability and survival on various surfaces where longest duration was reported for glass and plastic surface i.e. 84 h and 72 h, respectively while shortest time of droplet’s survival is 4 h in case of copper surface (Van Doremalen et al., 2020). Keeping in view its high rate of person to person transmission, series of actions have been recommended by WHO to restrict further spread of COVID-19 (Graham et al., 2013; Organization, 2020).

**Figure 1:**
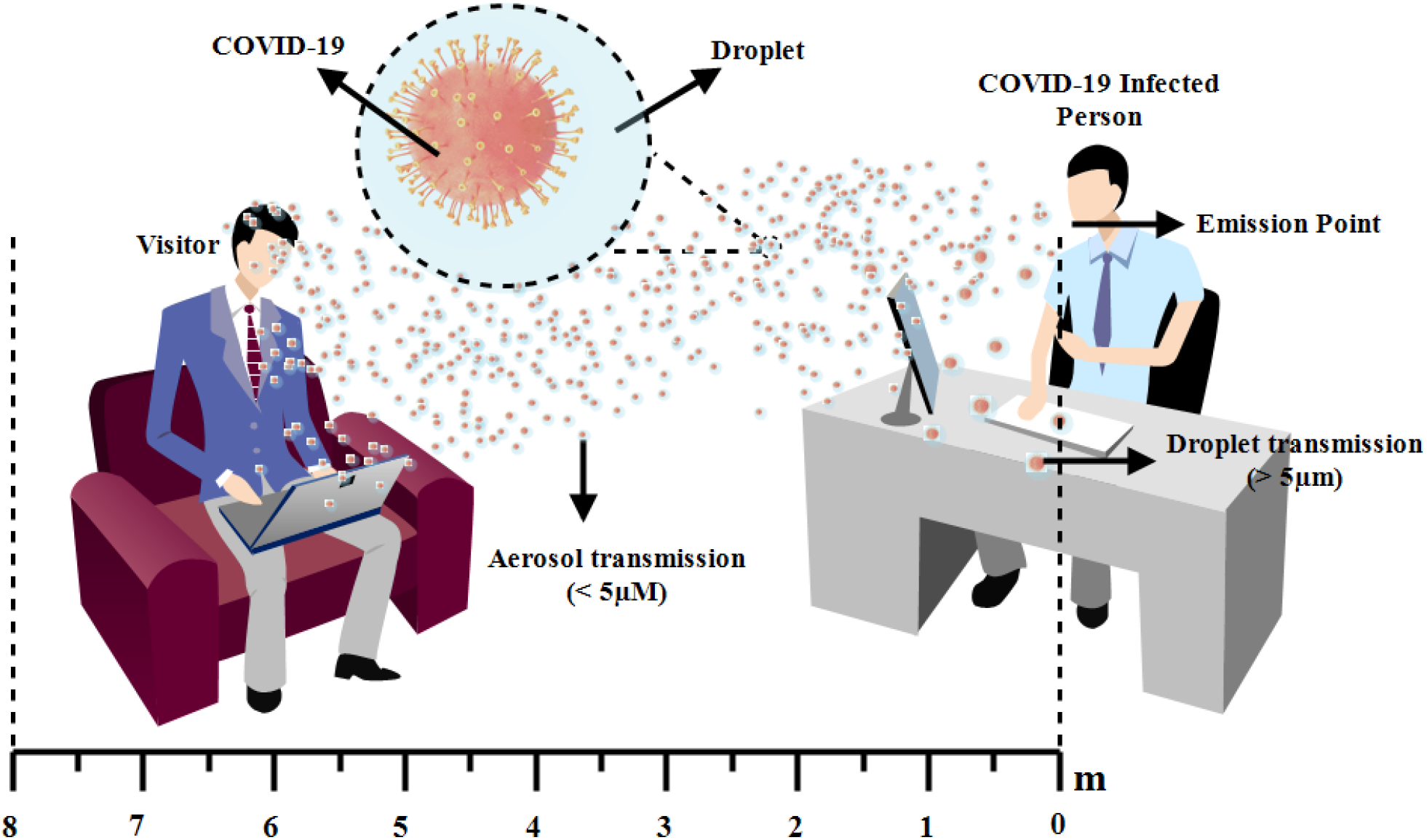
Trajectory of the transmission of COVID-19 by droplets and aerosols from an infected person.

Although spread of this viral infection is attributed to public mobility, person to person transmission and through respiratory droplets or direct contact but certain climatic factors also play pivotal role in spread of virus (Chen et al., 2020). Meteorological factors effecting the viability and spread of virus associated respiratory infections like SARS involve ambient temperature, humidity, population size and wind speed as reported by recent epidemiological studies (Dalziel et al., 2018; Ma et al., 2020; Tan et al., 2005). Stability of droplet and survival of coronavirus is dependent on air temperature as well as humidity (Chan et al., 2011). Recently, Tosepu et al., studied correlation of weather parameters and COVID-19 pandemic and reported positive linear correlation between average temperature and cases of COVID-19 in China and Indonesia (Tosepu et al., 2020; Zhu and Xie, 2020). Wang et al. reported effect of temperature on spread and mortality of COVID-19 while Metz and co-workers examined correlation of humidity with survival of virus (Metz and Finn, 2015; Wang et al., 2020).

Keeping in view, the significant correlation of climatological conditions with spread of COVID-19, the current study aimed to reveal correlation of COVID-19 pandemic and meteorological parameters including humidity, temperature, rainfall, wind speed and population size. Furthermore, ROC analysis was employed to investigate sensibility and sensitivity of population size, temperature average, temperature minimum, and wind speed on the transmission rate of the novel coronavirus in highly infected cities of Pakistan i.e. Karachi, Lahore and Peshawar to interlink relationship of their effects with COVID-19 mortality.

## 2. Materials and Methods

### 2.1 Data Collection

Data set of COVID-19 in Pakistan for the period of April 9 to June 9, 2020 was collected from the official website of National Institute of Health (https://www.nih.org.pk/) and daily confirmed cases of three highly infected cities (Lahore, Karachi and Peshawar) of Pakistan were gathered from online COVID-19 data archive (https://public.tableau.com/). The location and number of cases of highly infected cities including Lahore, Karachi and Peshawar are shown in Fig. 2A while, the weather data was taken from (https://weatherspark.com/) and (https://www.timeanddate.com/). The targeted climate variables are temperature minimum (°F), temperature maximum (°F), temperature average (°F), humidity (%), rainfall (mm) and windfall (mph). Population data of targeted cities were collect on the official website of Pakistan bureau of statistics (http://www.pbs.gov.pk/).

**Figure 2:**
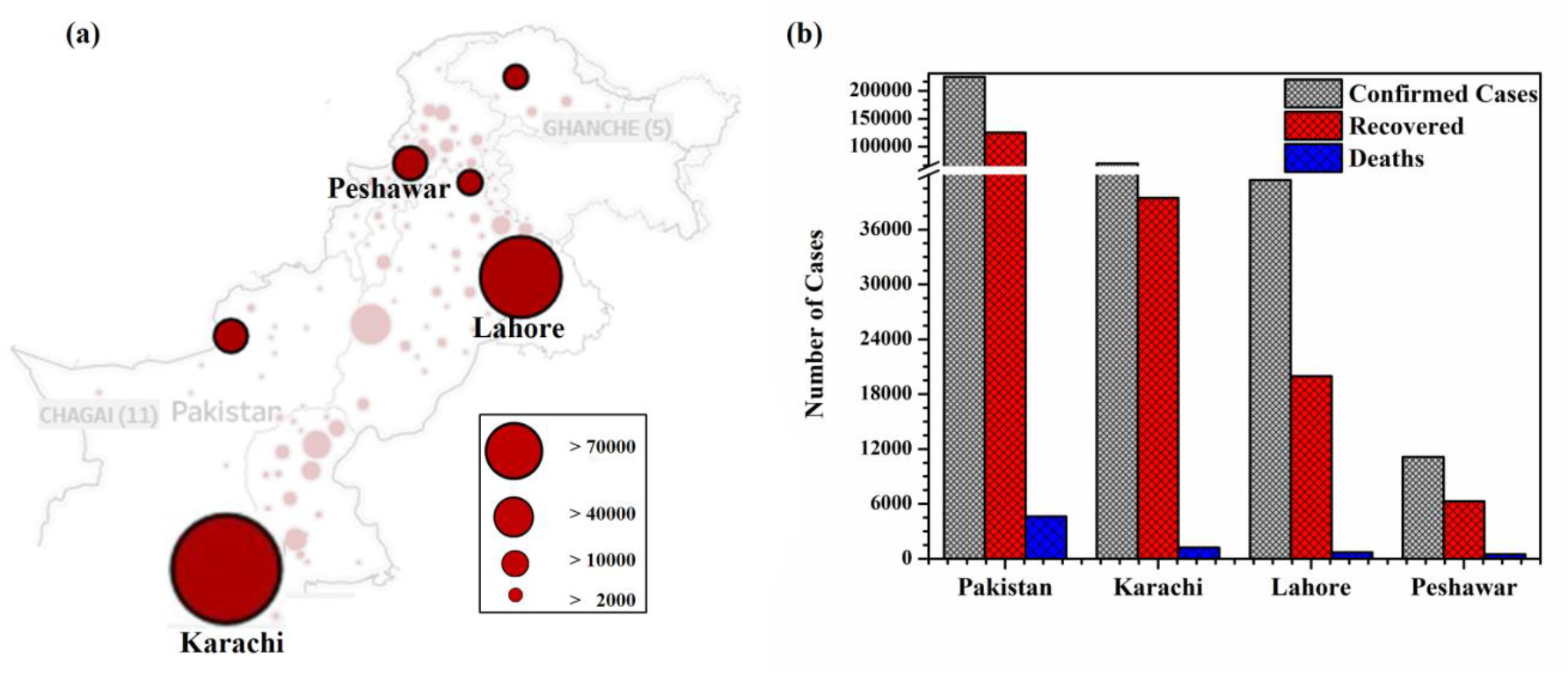
(a) COVID-19 epidemic in various cities of Pakistan and (b) numerical values for the total number of cases as on July 4, 2020.

### 2.2 Spearman’s correlation test

The collected data was analyzed by spearman’s rank correlation coefficient (*rs*) or Spearman’s rho (*ρ*) to determine the appropriate relationship between climatic variables and COVID-19 cases of targeted cities. It is similar to Pearson correlation coefficient and is non-parametric test that analyzes how well the association between two variables can be defined using a monotonic function. The correlation coefficient values of ±1 represents perfect degree of association between the two variables i.e. values closer to ±1 means stronger correlation while value near 0 means weaker correlation.

Given that the data used in this study are not normally distributed, it is appropriate to use correlation coefficients for the analyses that can be calculated via the following equation.

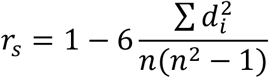

where n represents the number of alternatives, and di is the difference between the ranks of two parameters. All statistical analysis was performed using Microsoft excel 2010.

### 2.3 The analysis of receiver operating characteristics (ROC)

A probability curve known as receiver operating characteristic (ROC) curve has been widely employed for comparison between two operating characteristics i.e. true positive rate (TPR) or sensitivity and false positive rate (FPR) or specificity at different threshold settings (Hanley, 1989; Swets, 1979). Area under ROC curve an effective measure of accuracy is employed in current study to confirm correlation of climatic factors including temperature average (TA), temperature minimum (TM), wind speed (WS), and population size (PS) with transmission of COVID-19. GraphPad Prism software version 7.0 (GraphPad, Inc., San Diego, CA, USA, www.graphpad.com) is employed for ROC analysis while AUC was determined with 95% confidence intervals (CI). The AUC value for a perfect model is 1, worthless model is 0.5 and imperfect model is 0. The ROC curve is also referred as 1-specificity and sensitivity curve.

## 3. Results and Discussion

In Pakistan, first diagnosed case of COVID-19 has been confirmed on February 26, 2020 (Waris et al., 2020). A rapid increase in confirmed cases of COVID-19 were observed from March 16, 2020 onward, and on July 4, 2020 the total confirmed cases reached 225,282 as shown in Fig. 2B. The total number of confirmed cases, deaths and recoveries of three most infected cities such as Lahore Karachi and Peshawar of Pakistan until July 4, 2020 are also provided (Fig. 2B).

Fig. 3 shows the data of COVID-19 cases and climatic factors including temperature average, humidity and wind speed of selected cities. During this observation period (April 9, 2020 to June 9, 2020), total number of confirmed cases from selected cities were > 57000 with average daily number as 303. The average daily mean values of climatic factors considered in current study such as temperature minimum, temperature maximum, temperature average, rainfall, humidity average and wind speed were 73.3 °F, 92.8 °F, 83.0 °F, 17.2 mm, 53.6 % and 9.1 mph, respectively.

**Figure 3:**
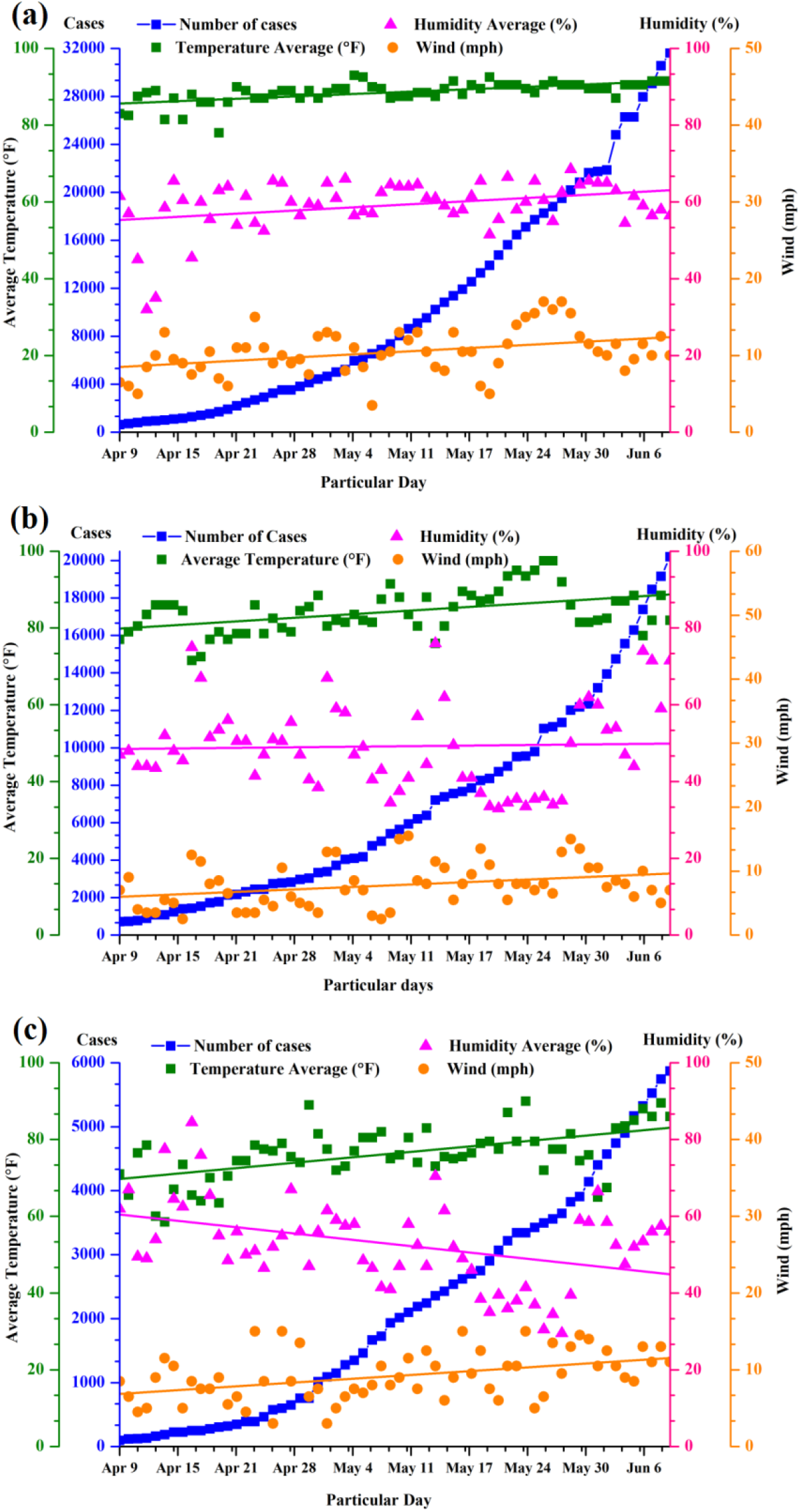
Daily confirmed cases of COVID-19 with climatic factors (Average temperature, wind speed and average humidity) in Pakistani cities (a) Karachi, (b) Lahore and (c) Peshawar from April 9, 2020 to June 9, 2020.

Table 1 summarizes the spearman’s correlation coefficient between daily COVID-19 confirmed cases and climatic variables along with population of selected cities. The result indicates significant correlation of confirmed cases with minimum temperature, average temperature and wind speed for each city while relative humidity is slightly correlated to the cases of Karachi. Finally, as expected, population of each city is strongly correlated with their number of cases. Among selected cities, Karachi is most infected by COVID-19, as assured by resulted statistical relationship of the number of cases with minimum temperature, average temperature, wind speed and population.

**Table 1:** Spearman’s correlation coefficient between daily COVID-19 confirmed cases and climatic variables along with population of selected cities in Pakistan.

| Sr. No | Climatic variables | Lahore | Karachi | Peshawar |
| --- | --- | --- | --- | --- |
| 1 | Temperature Minimum (°F) | 0.33* | 0.68* | 0.30* |
| 2 | Temperature Maximum (°F) | 0.21 | 0.11*** | 0.26** |
| 3 | Temperature Average (°F) | 0.37** | 0.56* | 0.35* |
| 4 | Rainfall (mm) | 0.19*** | -0.07 | -0.72* |
| 5 | Humidity average (%) | 0.07 | 0.10 | -0.19*** |
| 6 | Wind speed (mph) | 0.20 | 0.35* | 0.17*** |
| 7 | Population | 0.70* | 0.75* | 0.65* |

\* Correlation is significant at the 0.01, 0.05 and 0.1 level, respectively (2-tailed).

ROC analysis was originated in early 1950’s and provides an efficient method to evaluate accuracy of a test using intrinsic measures like sensitivity, and specificity. ROC curve (sensitivity versus 1-specificity plot) and area under ROC curve represents a reliable tool for performance measurement of test to (i) find out optimal cut off values, and (ii) comparison between alternative diagnostic tasks applied on same subject (Hajian-Tilaki, 2013; Hanley and McNeil, 1982; Hanley and McNeil, 1983). The ROC curve is utilized in current study for sensitivity and specificity analysis over three climatological parameters namely temperature average, temperature minimum and wind speed along with population size in highly effected cities of Pakistan. ROC curves for transmission rate of COVID-19 based on variation in various parameters in selected cities are depicted in Figure 4 where, the design variable is shown on horizontal axis and sensitivity of same variable is represented on vertical axis.

**Figure 4:**
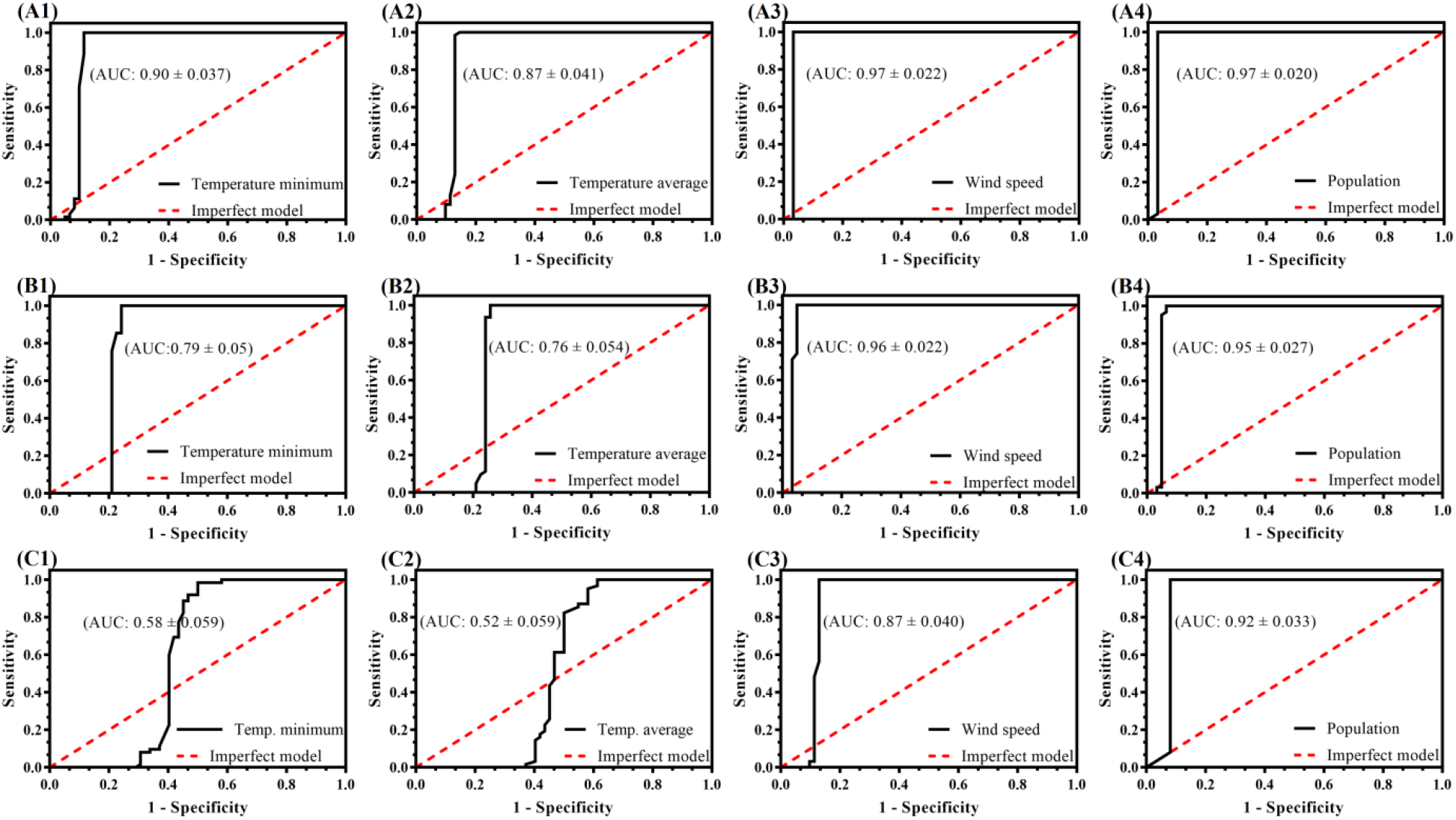
The ROC curve of the transmission rate of COVID-19 based on the various meteorological factors (TM, TA, and WS) and PS in Pakistani cities (a) Karachi, (b) Lahore and (c) Peshawar from April 9, 2020 to June 9, 2020. Specificity represents the true negative rate and sensitivity represents the true positive rate.

Figure 4 revealed that both WS and PS model with AUC value 0.9 have high sensitivity and can distinguish positive and negative classes for all selected cities. Similarly, TA and TM models generated for highly effected city i.e. Karachi with AUC value of 0.8 and 0.9, respectively also have good sensitivity and accurately distinguish both classes. In addition, TA and TM models for Lahore (second highly effected city) with AUC value 0.7 has moderate sensitivity while in case of Peshawar the generated TA and TM models have worst sensitivity (AUC: 0.5) as given in Table 2. The nonlinear relationship of TA, TM and PS parameters with number of COVID-19 infected people confirmed sensitivity and direct correlation of COVID-19 transmission with these parameters.

**Table 2:**
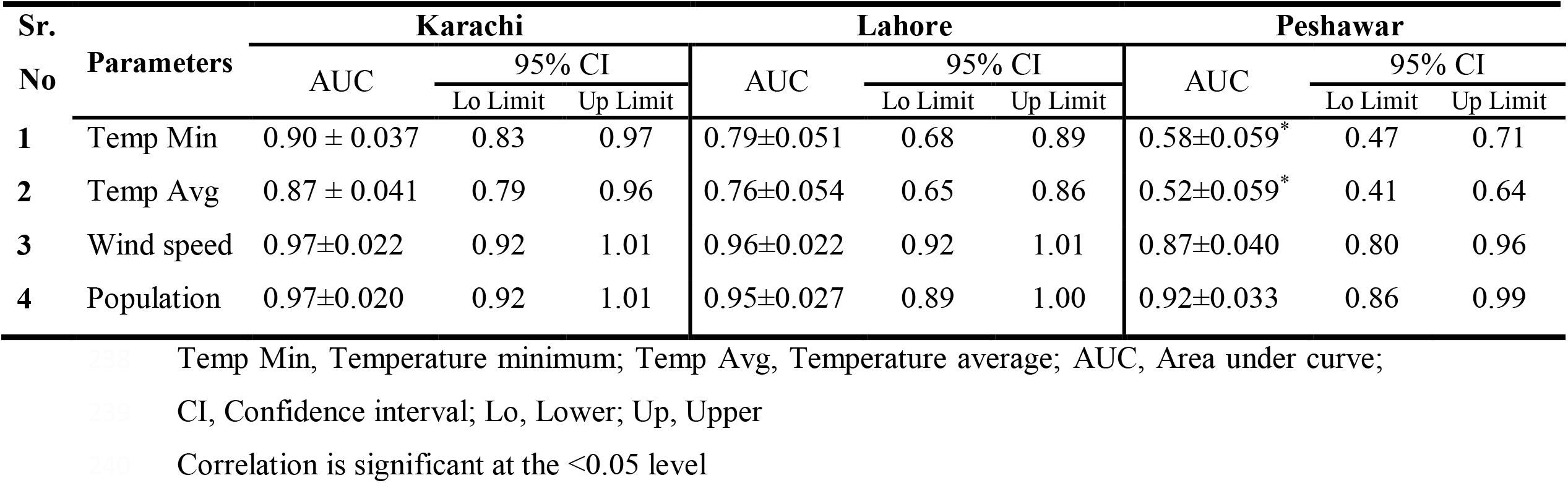
The area under curve of the transmission rate of COVID-19 based on the various meteorological factors and population of selected cities in Pakistan.

| Sr.<br>No | Parameters | Karachi |  |  | Lahore |  |  | Peshawar |  |  |
| --- | --- | --- | --- | --- | --- | --- | --- | --- | --- | --- |
|  |  | AUC | 95% CI |  | AUC | 95% CI |  | AUC | 95% CI |  |
|  |  |  | Lo Limit | Up Limit |  | Lo Limit | Up Limit |  | Lo Limit | Up Limit |
| 1 | Temp Min | 0.90 ± 0.037 | 0.83 | 0.97 | 0.79±0.051 | 0.68 | 0.89 | 0.58±0.059* | 0.47 | 0.71 |
| 2 | Temp Avg | 0.87 ± 0.041 | 0.79 | 0.96 | 0.76±0.054 | 0.65 | 0.86 | 0.52±0.059* | 0.41 | 0.64 |
| 3 | Wind speed | 0.97±0.022 | 0.92 | 1.01 | 0.96±0.022 | 0.92 | 1.01 | 0.87±0.040 | 0.80 | 0.96 |
| 4 | Population | 0.97±0.020 | 0.92 | 1.01 | 0.95±0.027 | 0.89 | 1.00 | 0.92±0.033 | 0.86 | 0.99 |

Temp Min, Temperature minimum; Temp Avg, Temperature average; AUC, Area under curve; CI, Confidence interval; Lo, Lower; Up, Upper Correlation is significant at the <0.05 level

In this study, we explore the statistical relationship between climatic variables and COVID-19 cases by spearman’s correlation coefficient method followed by ROC curves analysis to investigate the sensitivity of TA, TM, WS and PS on transmission rate of COVID-19 aerosol droplets in highly afflicted cities of Pakistan. Airborne transmission also termed as “droplet based transmission” of infectious agent (i.e. COVID-19) is responsible for unexpected escalation of this epidemic around the globe. Aerosols are liquid or solid particle suspension with virus encapsulated inside and pose high threat towards disease spread (Judson and Munster, 2019; Morawska, 2005). The viability and survival of these viral encapsulated droplets is effected by various environmental factors namely temperature variations, wind speed, humidity, and sunlight. Infectivity of virus (i.e. COVID-19; size< 100 nm) encapsulated aerosols depends mainly on stress caused by temperature along with other environmental factors (Kumar and Morawska, 2020; Marthi, 1994). Association of temperature with spread of viral particles has been extensively reported suggesting direct relation of temperature with viability and infectivity rate of different viral infections. Tan et al., reported higher rate of SARS-CoV cases in places of minimum and ambient temperature zone (Tan et al., 2005). Current study suggested that minimum and average air temperature facilitate the spread and transmission of this SARS-CoV-2 embedded inside aerosol particles and has positive correlation with the increasing number of COVID-19 cases. Our findings are in agreement with the previous reports of (Biswas et al., 2014; Harper, 1961; Shaman and Kohn, 2009; Van Doremalen et al., 2013) suggesting more survival rate of SARS-CoV, MERS-CoV and other influenza virus in low temperature zone and corresponding increase in viral pandemic. Best correlation of temperature minimum and temperature average with number of COVID-19 cases was observed in case of Karachi (highly effected city of Pakistan) where evaporation is an important physiochemical process that affect fate of COVID-19 droplets. Recent studies of Tosepu et al., 2020 support our findings that elucidated the significant correlation between average temperature and COVID-19 cases. Similarly, negative correlation observed in case of rainfall and average humidity in current study are also supported by previous studies of Tan et al 2010 and Metz & Finn et al indicating inverse relation of these variables with number of cases (Metz and Finn, 2015; Tan et al., 2005). Negative correlation observed in case of average humidity in current study is consistent with work reported by Chan et al., 2011 suggesting inverse relation among humidity and SARS coronavirus cases (Chan et al., 2011). Impact of climate on the number of COVID-19 cases in various countries has been reported previously by various research groups to explain the statistical interaction (Bashir et al., 2020; Şahin, 2020; Tosepu et al., 2020; Zhu and Xie, 2020). Furthermore, studies reported over COVID-19 epidemic from China also suggested strong correlation of coronavirus cases and various climatic parameters particularly, temperature and humidity (Sajadi et al., 2020).

Wind speed is another important climatic factor that facilitate the spread of infectious diseases like SARS-CoV, COVID-19 and influenza virus (Bashir et al., 2020; Yuan et al., 2006). Positive correlation of wind speed with COVID-19 spread is observed in current study suggesting role of wind for spread of COVID-19 aerosol particles to distance far from their origin. Furthermore, the ROC curve analysis (AUC value range 0.8 to 0.9) also showed good sensitivity of wind speed with number of COVID-19 cases in selected cities of Pakistan.

Population size (PS) is reported as major transmittability factor for COVID-19 and showed positive association with COVID-19 epidemic as depicted in Figure 2 while ROC curve analysis also confirmed sensitivity of PS towards spread of COVID-19. These findings are comparable to previous studies of Mehdi et al., that showed strong correlation of PS with number of COVID-19 cases (Jahangiri et al., 2020). Role of temperature (i.e. minimum and average), wind speed and population size on COVID-19 spread along with its survival rate and transmission risk on surfaces and air, respectively is well-documented. Therefore, sensitivity analysis of these parameters over transmission rate of COVID-19 in selected cities of Pakistan is crucial for government agencies to devise improved safety measures for survival of people. Owing to significance of PS parameter towards COVID-19 pandemic, more strict actions and control measures are suggested for highly populated cities to control devastating effects of COVID-19. Although our findings suggested strong correlation of COVID-19 epidemic and various meteorological parameters involving extensive data analysis from various cities of Pakistan. Still, current study has certain limitations as we did not consider other factors like personal hygeine, people mobility and endurance that need to be considered in further studies.

## Conclusions

Transmission of COVID-19 droplets and aerosols emitted by cough or sneeze of COVID-19 infected person is affected by various environmental factors along with population size. Current study investigated correlation of COVID-19 epidemic with different meteorological parameters particularly, temperature, wind speed, and population size of various cities of Pakistan. Unique characteristics of COVID-19 aerosols being more viable with long survival time in air pose more threat towards its spread. Our findings indicated significant correlation between temperature (i.e. TA and TM) and population size (PS) with COVID-19 pandemic. Furthermore, ROC curves were used to analyze the sensitivity of TA, TM, WS and PS on transmission rate of COVID-19 aerosol particles in highly afflicted cities of Pakistan. The results showed non-linear relationship of TA, TM and PS with number of people effected from COVID-19 in selected cities of Pakistan and sensitivity of all selected parameters with COVID-19 transmission. Results of current study suggest temperature variation and population size as important factors affecting the COVID-19 epidemic and can be used by government authorities to devise policy to suppress epidemic in Pakistan.

## Data Availability

Data used for current study has been provided in manuscript.

## Acknowledgement

The authors thank Dr. Hafiz Muhammad Adnan Hammed and Muhammad Zahid for scientific discussions. Junaid Haider is thankful to “CAS-TWAS President’s Fellowship for International PhD Students”.

## Author contribution

IS, AS and JH designed concept of current study, arranged data from various resources and applied statistical analysis. SN, IS and AH analyzed data, compiled information and wrote manuscript. RMA, HRS and IMK contributed literature, and analyzed data. MI reviewed manuscript drafts, finalized formatting and supervised the study. All authors have given approval to the final version of the manuscript.

## References

Ali, I., 2020 Pakistan confirms first two cases of coronavirus, govt says “no need to panic”. DAWN, 2020.

Bashir, M.F., Ma, B., Komal, B., Bashir, M.A., Tan, D., Bashir, M., 2020. Correlation between climate indicators and COVID-19 pandemic in New York, USA. Sci. Total. Environ. 728, 138835.

Biswas, P.K., Islam, M.Z., Debnath, N.C., Yamage, M., 2014. Modeling and roles of meteorological factors in outbreaks of highly pathogenic avian influenza H5N1. PloS one 9, e98471.

Chan, K., Peiris, J., Lam, S., Poon, L., Yuen, K., Seto, W., 2011. The effects of temperature and relative humidity on the viability of the SARS coronavirus. Adv. Virol. 2011, 734690.

Chen, B., Liang, H., Yuan, X., Hu, Y., Xu, M., Zhao, Y., et al. Roles of meteorological conditions in COVID-19 transmission on a worldwide scale. MedRxiv 2020.

Dalziel, B.D., Kissler, S., Gog, J.R., Viboud, C., Bjørnstad, O.N., Metcalf, C.J.E., et al. 2018. Urbanization and humidity shape the intensity of influenza epidemics in US cities. Science 362, 75–79.

Graham, R.L., Donaldson, E.F., Baric, R.S., 2013. A decade after SARS: strategies for controlling emerging coronaviruses. Nat. Rev. Microbiol. 11, 836–848.

Grayson, S.A., Griffiths, P.S., Perez, M.K., Piedimonte, G., 2017 Detection of airborne respiratory syncytial virus in a pediatric acute care clinic. Pediatr. pulmonol. 52, 684–688.

Guan, W-j., Ni, Z-y., Hu, Y., Liang, W-h., Ou, C-q., He, J-x., et al., 2020. Clinical characteristics of coronavirus disease 2019 in China. N. Engl. J. Med. 382, 1708–1720.

Hajian-Tilaki, K., 2013. Receiver operating characteristic (ROC) curve analysis for medical diagnostic test evaluation. Caspian. J. Intern. Med. 4, 627.

Hanley, J.A., 1989. Receiver operating characteristic (ROC) methodology: the state of the art. Crit. Rev. Diagn. Imaging. 29, 307–335.

Hanley, J.A., McNeil, B.J., 1982. The meaning and use of the area under a receiver operating characteristic (ROC) curve. Radiology 143, 29–36.

Hanley, J.A., McNeil, B.J., 1983. A method of comparing the areas under receiver operating characteristic curves derived from the same cases. Radiology 148, 839–843.

Harper, G., 1961. Airborne micro-organisms: survival tests with four viruses. Epidemiol. Infect. 59, 479–486.

Huang, C., Wang, Y., Li, X., Ren, L., Zhao, J., Hu, Y., et al., 2020. Clinical features of patients infected with 2019 novel coronavirus in Wuhan, China. The lancet 395, 497–506.

Jahangiri, M., Jahangiri, M., Najafgholipour, M., 2020. The sensitivity and specificity analyses of ambient temperature and population size on the transmission rate of the novel coronavirus (COVID-19) in different provinces of Iran. Sci. Total. Environ, 138872.

Jayaweera, M., Perera, H., Gunawardana, B., Manatunge, J., 2020. Transmission of COVID-19 virus by droplets and aerosols: A critical review on the unresolved dichotomy. Environ. Res, 109819.

Judson, S.D., Munster, V.J., 2019. Nosocomial transmission of emerging viruses via aerosol-generating medical procedures. Viruses 11, 940.

Kumar, P., Morawska, L., 2020. Could fighting airborne transmission be the next line of defence against COVID-19 spread? City and Environment Interactions, 100033.

Li, Q., Guan, X., Wu, P., Wang, X., Zhou, L., Tong, Y., et al., 2020. Early transmission dynamics in Wuhan, China, of novel coronavirus–infected pneumonia. N. Engl. J. Med. 382, 1199–1207.

Lipsitch, M., Swerdlow, D.L., Finelli, L., 2020. Defining the epidemiology of Covid-19—studies needed. N. Engl. J. Med. 382, 1194–1196.

Liu, L., Wei, J., Li, Y., Ooi, A., 2017. Evaporation and dispersion of respiratory droplets from coughing. Indoor Air 27, 179–190.

Ma, Y., Zhao, Y., Liu, J., He, X., Wang, B., Fu, S., et al. 2020. Effects of temperature variation and humidity on the death of COVID-19 in Wuhan, China. Sci. Total. Environ, 138226.

Marthi, B., 1994. Resuscitation of microbial bioaerosols. Atmospheric microbial aerosols. Springer, 192–225.

Metz, J.A., Finn, A., 2015. Influenza and humidity–Why a bit more damp may be good for you!. J. Infect. 71, S54–S58.

Morawska, L., 2005. Droplet fate in indoor environments, or can we prevent the spread of infection? Proceedings of Indoor Air 2005: the 10th International Conference on Indoor Air Quality and Climate. Springer, 2005, 9–23.

Nicas, M., Nazaroff, W.W., Hubbard, A., 2005. Toward understanding the risk of secondary airborne infection: emission of respirable pathogens. J. Occup. Environ. Hyg. 2, 143–154.

Noreen, N., Dil, S., Niazi, S.K., Naveed, I., Khan, N.U., Khan, F.K., et al., 2020. Coronavirus disease (COVID-19) Pandemic and Pakistan; Limitations and Gaps.

World Health Organization (WHO) 2020. Critical preparedness, readiness and response actions for COVID-19: interim guidance, 19 March 2020.

Remuzzi, A., Remuzzi, G., COVID-19 and Italy: what next? The Lancet 2020.

Rothan, H.A., Byrareddy, S.N., 2020. The epidemiology and pathogenesis of coronavirus disease (COVID-19) outbreak. J. Autoimmun. 109, 102433.

Şahin, M., 2020. Impact of weather on COVID-19 pandemic in Turkey. Sci. Total. Environ. 728, 138810.

Sajadi, M.M., Habibzadeh, P., Vintzileos, A., Shokouhi, S., Miralles-Wilhelm, F., Amoroso, A., 2020. Temperature and latitude analysis to predict potential spread and seasonality for COVID-19. Available at SSRN 3550308.

Saqlain, M., Munir, M.M., Ahmed, A., Tahir, A.H., Kamran, S., 2020. Is Pakistan prepared to tackle the coronavirus epidemic? Drugs. Ther. Perspect, 1-2.

Shaman, J., Kohn, M., 2009. Absolute humidity modulates influenza survival, transmission, and seasonality. Proc. Natl. Acad. Sci. 106, 3243–3248.

Song, F., Shi, N., Shan, F., Zhang, Z., Shen, J., Lu, H., et al. 2020. Emerging 2019 novel coronavirus (2019-nCoV) pneumonia. Radiology 295, 210–217.

Swets, J.A., 1979. ROC analysis applied to the evaluation of medical imaging techniques. Invest. Radiol. 14, 109–121.

Tan, J., Mu, L., Huang, J., Yu, S., Chen, B., Yin, J., 2005. An initial investigation of the association between the SARS outbreak and weather: with the view of the environmental temperature and its variation. J. Epidemiol. Commun. H. 59, 186–192.

Tellier, R., 2009. Aerosol transmission of influenza A virus: a review of new studies. J. R. Soc. Interface 6, S783–S790.

Tosepu, R., Gunawan, J., Effendy, D.S., Lestari, H., Bahar, H., Asfian, P., 2020. Correlation between weather and Covid-19 pandemic in Jakarta, Indonesia. Sci. Total. Environ. 725, 138436.

Van Doremalen, N., Bushmaker, T., Morris, D.H., Holbrook, M.G., Gamble, A., Williamson, B.N., et al., 2020. Aerosol and surface stability of SARS-CoV-2 as compared with SARS-CoV-1. N. Engl. J. Med. 382, 1564–1567.

Van Doremalen, N., Bushmaker, T., Munster, V., 2013. Stability of Middle East respiratory syndrome coronavirus (MERS-CoV) under different environmental conditions. Eurosurveillance 18, 20590.

Wang, M., Jiang, A., Gong, L., Luo, L., Guo, W., Li, C., et al., 2020. Temperature significant change COVID-19 Transmission in 429 cities. MedRxiv 2020.

Waris, A., Khan, A.U., Ali, M., Ali, A., Baset, A., 2020. COVID-19 outbreak: current scenario of Pakistan. New. Microbes. New. Infect, 100681.

Xie, X., Li, Y., Chwang, A., Ho, P., Seto, W., 2007. How far droplets can move in indoor environments–revisiting the Wells evaporation–falling curve. Indoor air 17, 211–225.

Yang, P., Wang, X., 2020. COVID-19: a new challenge for human beings. Cell. Mol. Immunol. 17, 555–557.

Yuan, J., Yun, H., Lan, W., Wang, W., Sullivan, S.G., Jia, S., et al., 2006. A climatologic investigation of the SARS-CoV outbreak in Beijing, China. Am. J. Infect. Control 34, 234–236.

Zhu, H., Wei, L., Niu, P., 2020. The novel coronavirus outbreak in Wuhan, China. Global health research and policy 5, 1–3.

Zhu, Y., Xie, J., 2020. Association between ambient temperature and COVID-19 infection in 122 cities from China. Sci. Total. Environ. 724, 138201.

